# Socioeconomic inequities in care experienced by women with breast cancer in England: An intersectional cross-sectional study

**DOI:** 10.1101/2023.12.13.23299922

**Authors:** Mar Estupiñán Fdez de Mesa, Afrodita Marcu, Emma Ream, Katriina L Whitaker

**Affiliations:** School of Health Sciences, University of Surrey, Research Park, Guildford, Surrey, UK, GU2 7YH

## Abstract

**Purpose:** Guided by the intersectionality framework, we examined the differential in breast cancer care experience across population subgroups in England.

**Methods:** Secondary data analysis using the 2017/2018 English National Cancer Patient Experience Survey (NCPES). We applied disaggregated descriptive statistics (mean, standard errors, 95% confidence interval) to analyse 26,030 responses from female breast cancer patients to a question relating to overall care experience categorised by age, ethnicity, and sexual orientation in their intersection with deprivation status. We then applied multivariable logistic regression (odds ratios, 95% confidence intervals) to ascertain the relationship of reporting a positive care experience adjusting for patient, clinical, and trust-level factors.

**Results:** Poorer breast cancer care experience was mostly reported by the most deprived younger and minoritised ethnic groups. Similar findings were observed in adjusted multivariable analyses. Younger respondents were less likely than older patients to rate their care favourably. Pakistani, Indian, Chinese, and Black African women were less likely than White British women to rate their care favourably. Respondents from the most socioeconomic deprived backgrounds were less likely than the most affluent ones to rate their care favourably.

**Conclusion:** There is evidence of inequity in overall cancer care experience among female breast cancer patients in England, particularly among women living at the specific intersection of age, ethnicity and socioeconomic position. Future research is necessary to understand the mechanisms underlying breast cancer inequities. Policymakers, commissioners, and providers should consider the existence of multiple forms of marginalization to inform improvement initiatives targeting patients at higher risk of vulnerability.

## Introduction

Cancer is a growing global issue and a public health priority that imposes a profound burden on historically marginalised populations (IARC, 2019). Within this scenario, breast cancer requires specific attention. In 2020, breast cancer became the world’s most prevalent cancer overtaking lung cancer, with over the past 5 years an estimated 7.8 million women living with the disease (WHO, 2021a). Breast cancer is the first or second leading cause of female deaths in 95% countries (WHO, 2023). This has a ripple effect in families, particularly for children. Recent evidence suggests that in 2020 approximately 1 million children were orphaned as a consequence of cancer through an estimated 4.4 million women dying from the disease, 25% of which were due to breast cancer (Ginsburg et al., 2023, WHO, 2023).

Globally, collaborative efforts are being made to reduce breast cancer mortality and improve cancer care and quality of life (WHO, 2021b, WHO, 2023). A central action to achieve this goal is to improve the management and delivery of breast cancer care. Patient care experience has been positively associated with clinical effectiveness and patient safety (Cathal et al., 2013), and it has become a key measure to improve cancer services (NHS, 2023). However, evidence shows that globally systematic differences in patient care experience exist based on gender (Wessels et al., 2010), and particularly among people of colour (Pinder et al., 2016, Saunders et al., 2015, Trenchard et al., 2016). To understand these differences, population health research has commonly homogenised groups assuming that group membership (e.g., ethnicity) is linked to shared experiences, culture, and beliefs (Darko, 2023). This approach is problematic because it essentialises categories such as gender (for instance, it considers all women have the same experience, views, and priorities regardless of their identities and social backgrounds)(Hankivsky et al., 2010), and often fails to recognise that heterogeneity of experience exists between and within ethnic groups (Sayani, 2017). Further, examining social dimensions in isolation only provides a partial picture of cancer inequities. Instead, researchers are encouraged to focus on how socio-historical processes, intertwine with structural factors, to influence cancer outcomes (Kagan, 2014, Williams et al., 2012).

One way to address this challenge is to apply the intersectionality framework. Rooted in the Black Feminist movement (Collins, 2008, Combahee River Collective, 1977), intersectionality has been instrumental in challenging the conventional linear thinking that homogenises groups and prioritises mono-categorical lenses as the baseline to examine inequities (Hankivsky and Christoffersen, 2008). Rather, through intersectional lenses, socially-constructed identities (e.g., gender, ethnicity), occurring within broader contexts (e.g., culture, religion) and power structures (e.g. racism, discrimination) are seen to operate simultaneously to create privileges or challenges at the point they intersect (Hankivsky et al., 2010, Shannon et al., 2022a). It is at this intersecting social location where it is understood that unique social and health inequities arise (Sayani, 2017). For instance, evidence suggests that inequities in breast cancer examination among women in India emerge as a result of the intersecting effects of age, marital status, employment, religion, and place of residence (Negi and Nambiar, 2021). Similarly, delays in breast cancer care for African-American sexual minority women are related to the systems of oppression they experience (racism and stigma) as a result of being located at the intersection of race/ethnicity and sexual orientation (Poteat et al., 2021).

In England, care experience is collected annually through the National Cancer Patient Experience Survey (NCPES) (NHS England, 2022). Analyses of the NCPES survey have shown that women persistently rate their cancer experience (all tumours) less favourably than men (Bone et al., 2014, Saunders et al., 2015). In addition, younger patients and minority ethnic groups are more likely to rate their care less favourably than their older and White British counterparts, respectively (Bone et al., 2014, El Turabi et al., 2013, Pinder et al., 2016, Trenchard et al., 2016). Compared to all groups of people living with cancer, those with breast cancer are more likely to rate their care experience positively (Saunders et al., 2015); however, little is known about the differences in care experience among female breast cancer patients. Moreover, NCPES research has focused on individual social dimensions, and little is known about the experience of patients living at the intersection of multiple dimensions of inequity. To cover this gap, and guided by the intersectionality framework, our aim was to examine intracategorical differences of care experience among women with breast cancer living at the intersection of socioeconomic position and age, ethnicity, and sexual orientation. The research was framed in the historical, cultural, and societal context where women with breast cancer live and have access to universal National Health Service (NHS) in the UK.

## Methods

### Data source

Secondary data analysis was performed using publicly available, anonymous NCPES data available for non-for-profit research from the UK Data Service (UK Data Service). The survey is sent to all adults (aged 16 and over) with a confirmed primary diagnosis of cancer who attended treatment in an NHS Hospital in England (inpatient or outpatient). Non-respondents are followed up with two reminders. The survey contains 59 multiple-choice questions relative to cancer care experience and associated demographic data. Evidence shows that minority ethnic groups are consistently under-represented in the NCPES survey (Alessy et al., 2019). Hence, we used two consecutive, identical annual NCPES surveys (2017, 2018) to address the under-representation of minoritised groups and to increase statistical power. The datasets were anonymised at source, therefore ethical approval was not required to conduct secondary analysis of these national data.

In England, care experience is used alongside clinical effectiveness and patient safety to monitor and improve cancer services (NHS England, 2022). Previous NCPES studies have focused on understanding differences in care experience among minority ethnic groups (Pinder et al., 2016), patients’ involvement in decision-making (El Turabi et al., 2013), or research participation (Mc Grath-Lone et al., 2015). Overall care experience provides a useful insight of how patients perceive the quality of services they receive (Prakash et al., 2020). In this study we were interested to explore breast cancer patients’ care experience (dependent variable) based on full valid data to question *Q59 Overall, how would you rate your care?* The surveys collected overall care experience using a Likert scale (0 – very poor-to 10 –very good). We used this continuous variable for descriptive analysis. To compare our findings with previous research (Saunders et al., 2015, Trenchard et al., 2016), we subsequently recorded overall ratings into a binary variable (positive/negative).

Patient characteristics were collected from the associated demographic information on cancer patients and English NHS Trusts (i.e., hospitals where patients attended) included in the NCPES surveys. The eligible population was women (female hospital record) with a primary diagnosis of breast cancer (n=26,030) (Figure 1). Age was categorised into six groups (16-34, 35-44; 45-54; 55-64; 65-74; ≥75). The 65-74 category was used as the reference group as this was the largest group of participants. Self-reported ethnicity^1^ was the chosen measure for ethnicity because it is considered to be ‘gold standard’(Saunders et al., 2013). A broad six-group (White British, Other White, Asian, Black, Mixed, and Other ethnicity) and 13 sub-groups of UK ethnic classifications (ONS, 2022) were used in the analysis. Socioeconomic position was analysed using the Indices of Multiple Deprivation (IMD) quintiles (values 1 to 5, 1=most deprived) included in the NCPES surveys. The IMD is the official composite measure of area-level deprivation in England (2019). Self-reported sexual orientation was recoded into a binary variable (i.e., heterosexual, sexual minority groups). Only patients who were residents in England were included; non-resident patients who were referred to English NHS Trusts for treatment were excluded (n=3). Clinical factors included time since first treatment (<1year, 1-5 years, >5 years) and patient classification (day case, inpatient). Comorbidities were excluded from the analysis due to the high proportion of missing data (>5%). Hospitals where patients attended, seven NHS regions (NHS England), and type of hospital (teaching/others) were used as Trust-level factors.

**Figure 1.**
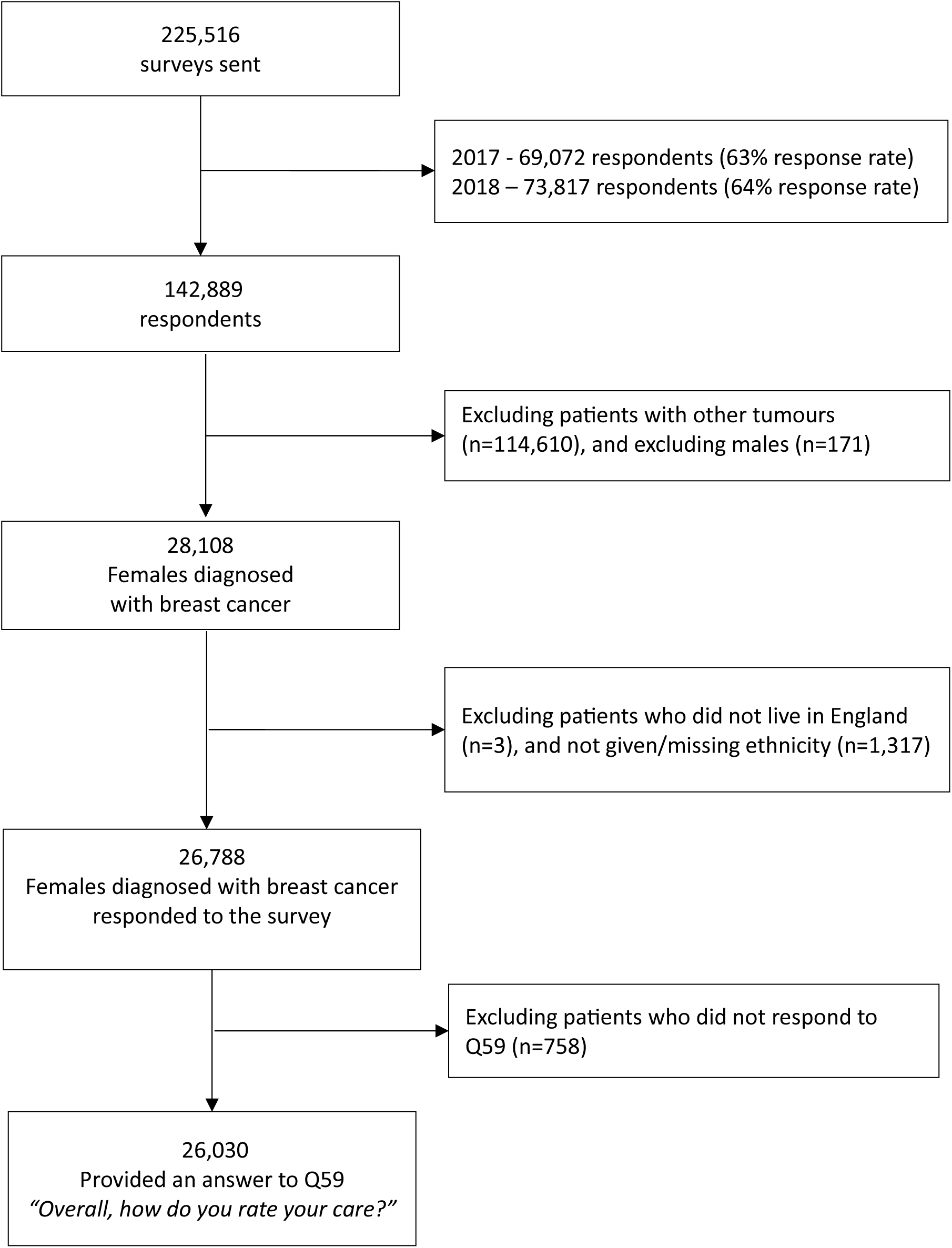
Flow diagram of the number of respondents included in the analysis. Number of 2017/2018 NCPES respondents included at each stage of the analysis.

### Statistical analysis

We sought to examine differential in the care experienced in relation to age, ethnicity, and sexual orientation among different socioeconomic groups using double disaggregation (Negi and Nambiar, 2021). Guided by quantitative intersectional analysis (Bauer, 2021, Bauer et al., 2021), we categorised each of the three dimensions (age, ethnicity, and sexual orientation) by deprivation quintile. First, we performed descriptive analysis to summarise respondents’ characteristics (frequency/percentage) (Table 1), followed by descriptive mean, standard errors and 95% confidence intervals of breast cancer patients’ care experience disaggregated by three dimensions of inequity and their intersections with deprivation status (Table 2). Chi square tests were used to find the association between positive care experience and selected dimensions. This analysis was supplemented by univariate logistic regression (Table 3) to examine the relationship between overall care experience and patient, clinical, and trust characteristics (as assessed by Q59) (model 1), followed by multivariable logistic regression to adjust for potential confounders added sequentially (patient factors (model 2) and clinical factors (model 3)). After controlling for fixed effects, mixed effects analysis suggested there was no variation for the random effect of clustering by trust factors among respondents. Hence multivariate logistic regression was chosen (model 4). Findings from regression logistic analyses were reported using Odds Ratios (ORs) and 95% Confidence Intervals (CIs); p-values <0.05 were deemed significant. All analyses were performed using SPSS Statistics 28.0.1 software.

**Table 1.**
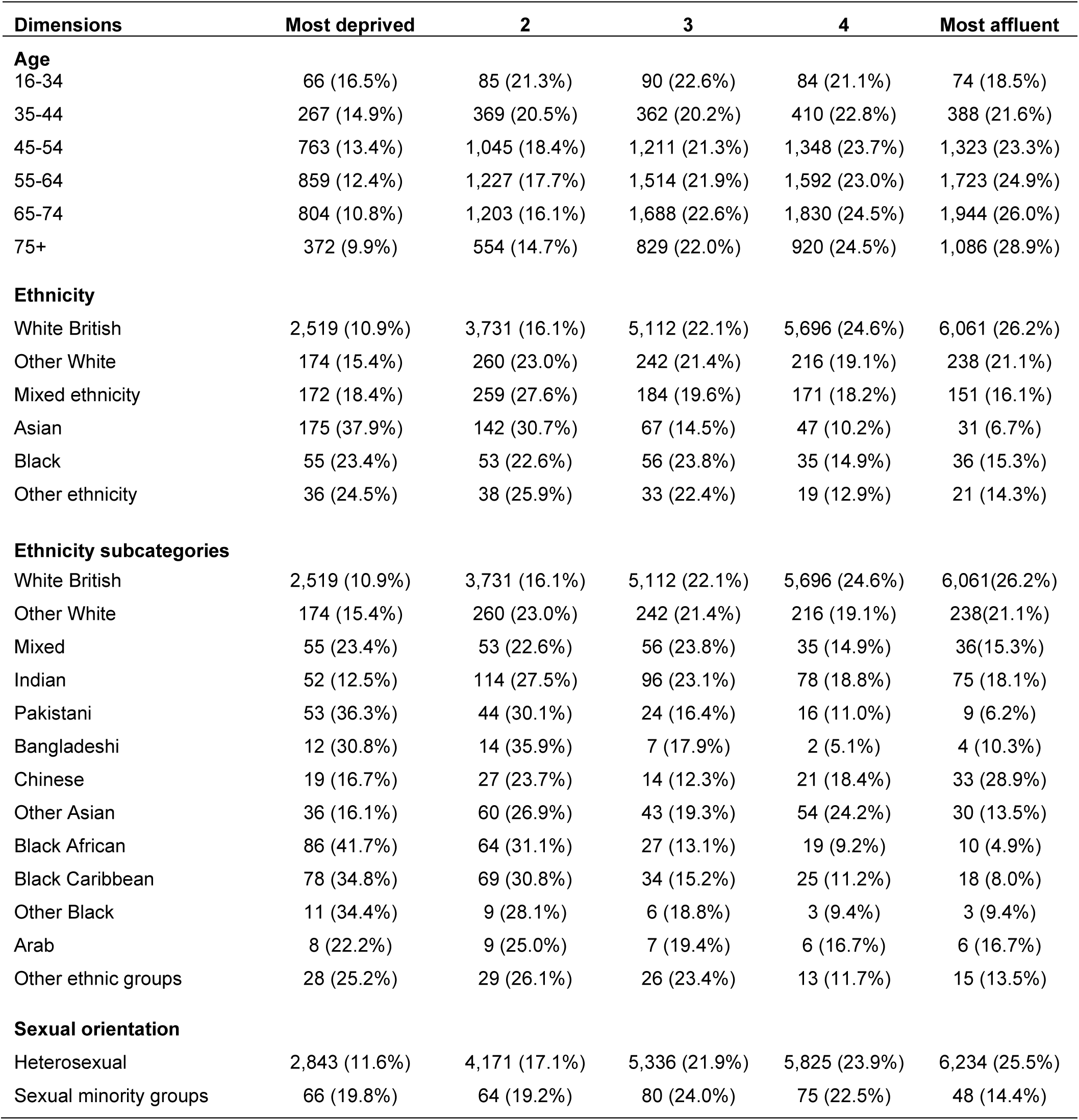
Demographic characteristics of respondents in the survey by deprivation quintiles represented by N (%), 2017/2018 NCPES.

**Table 2.** Descriptive analysis and summary measures for overall positive care experience using 2017/2018 NCPES.

| **Dimensions** | **Most deprived** | **2** | **3** | **4** | **Most affluent** |
| --- | --- | --- | --- | --- | --- |
| **Age** |  |  |  |  |  |
| 16-34 | 8.59 (8.12-9.06) | 8.39 (8.05-8.72) | 8.77 (8.50-9.03) | 8.67 (8.34-8.99) | 8.72 (8.33-9.10) |
| 35-44 | 8.50 (8.30-8.69) | 8.48 (8.32-8.64) | 8.64 (8.49-8.78) | 8.69 (8.57-8.82) | 8.60 (8.46-8.74) |
| 45-54 | 8.69 (8.58-8.80) | 8.76 (8.67-8.84) | 8.75 (8.67-8.83) | 8.73 (8.65-8.80) | 8.78 (8.70-8.85) |
| 55-64 | 8.88 (8.78-8.98) | 8.86 (8.78-8.93) | 8.89 (8.83-8.96) | 8.87 (8.81-8.94) | 8.90 (8.84-8.96) |
| 65-74 | 9.00 (8.91-9.10) | 9.01 (8.94-9.08) | 8.93 (8.87-8.99) | 8.96 (8.90-9.01) | 8.97 (8.91-9.02) |
| 75+ | 8.98 (8.85-9.11) | 8.98 (8.87-9.09) | 9.04 (8.96-9.13) | 8.92 (8.84-9.00) | 8.99 (8.92-9.06) |
| **Ethnicity** |  |  |  |  |  |
| White British | 8.92 (8.87-8.98) | 8.94 (8.89-8.98) | 8.92 (8.89-8.96) | 8.89 (8.85-8.92) | 8.91 (8.87-8.94) |
| Other White | 8.65 (8.41-8.89) | 8.55 (8.36-8.74) | 8.71 (8.54-8.88) | 8.64 (8.45-8.84) | 8.77 (8.59-8.95) |
| Mixed ethnicity | 8.64 (8.31-8.97) | 8.43 (8.08-8.79) | 8.63 (8.26-8.99) | 8.54 (8.08-9.00) | 8.81 (8.26-9.35) |
| Asian | 8.40 (8.12-8.67) | 8.31 (8.12-8.49) | 8.27 (8.06-8.49) | 8.32 (8.08-8.56) | 8.42 (8.16-8.69) |
| Black | 8.43 (8.22-8.64) | 8.42 (8.22-8.63) | 8.03 (7.59-8.47) | 8.64 (8.25-9.03) | 8.97 (8.60-9.34) |
| Other ethnicity | 8.44 (7.78-9.11) | 8.21 (7.69-8.74) | 8.48 (7.97-9.00) | 8.42 (7.44-9.40) | 8.95 (8.53-9.37) |
| **Ethnicity subcategories** |  |  |  |  |  |
| White British | 8.92 (8.87-8.98) | 8.94 (8.89-8.98) | 8.92 (8.89-8.96) | 8.89 (8.85-8.92) | 8.91 (8.87-8.94) |
| Other White | 8.65 (8.41-8.89) | 8.55 (8.36-8.74) | 8.71 (8.54-8.88) | 8.64 (8.45-8.84) | 8.77 (8.59-8.95) |
| Mixed | 8.64 (8.31-8.97) | 8.43 (8.08-8.79) | 8.63 (8.26-8.99) | 8.54 (8.08-9.00) | 8.81 (8.26-9.35) |
| Indian | 8.04 (7.34-8.74) | 8.35 (8.07-8.63) | 8.17 (7.86-8.47) | 8.21 (7.84-8.57) | 8.23 (7.80-8.65) |
| Pakistani | 8.45 (8.08-8.83) | 8.41 (7.91-8.91) | 7.96 (7.38-8.54) | 8.69 (7.92-9.46) | 9.44 (8.89-10.0) |
| Bangladeshi | 8.75 (8.14-9.36) | 8.57 (7.67-9.47) | 9.00 (7.59-10.41) | 8.00 (-4.71-0.71) | 9.25 (6.86-1.64) |
| Chinese | 8.47 (7.93-9.02) | 7.89 (7.28-8.49) | 7.79 (7.10-8.47) | 8.00 (7.36-8.64) | 8.42 (7.92-8.93) |
| Other Asian | 8.67 (8.11-9.23) | 8.27 (7.90-8.64) | 8.72 (8.27-9.17) | 8.52 (8.09-8.95) | 8.50 (7.90-9.10) |
| Black African | 8.34 (8.02-8.65) | 8.44 (8.12-8.75) | 7.81 (7.03-8.60) | 8.53 (7.80-9.25) | 8.90 (8.19-9.61) |
| Black Caribbean | 8.59 (8.27-8.91) | 8.42 (8.12-8.72) | 8.12 (7.49-8.75) | 8.68 (8.15-9.21) | 8.94 (8.42-9.47) |
| Other Black | 8.00 (7.33-8.67) | 8.33 (7.32-9.35) | 8.50 (7.40-9.60) | 9.00 (9.00-9.00) | 9.33 (6.46-12.20) |
| Arab | 7.50 (5.55-9.45) | 7.44 (6.11-8.78) | 8.29 (7.83-8.74) | 9.33 (8.48-10.19) | 9.00 (8.06-9.94) |
| Other ethnic groups | 8.71 (8.01-9.42) | 8.45 (7.88-9.02) | 8.54 (7.88-9.20) | 8.00 (6.60-9.40) | 8.93 (8.40-9.47) |
| **Sexual orientation** |  |  |  |  |  |
| Heterosexual | 8.85 (8.79-8.90) | 8.87 (8.83-8.91) | 8.89 (8.85-8.92) | 8.87 (8.84-8.90) | 8.89 (8.86-8.92) |
| Sexual minority groups | 8.86 (8.53-9.19) | 8.16 (7.74-8.58) | 8.86 (8.60-9.13) | 8.83 (8.48-9.18) | 8.75 (8.29-9.21) |

All subgroups, except sexual orientation, were statistically significant at $p < 0.05$

**Table 3.**
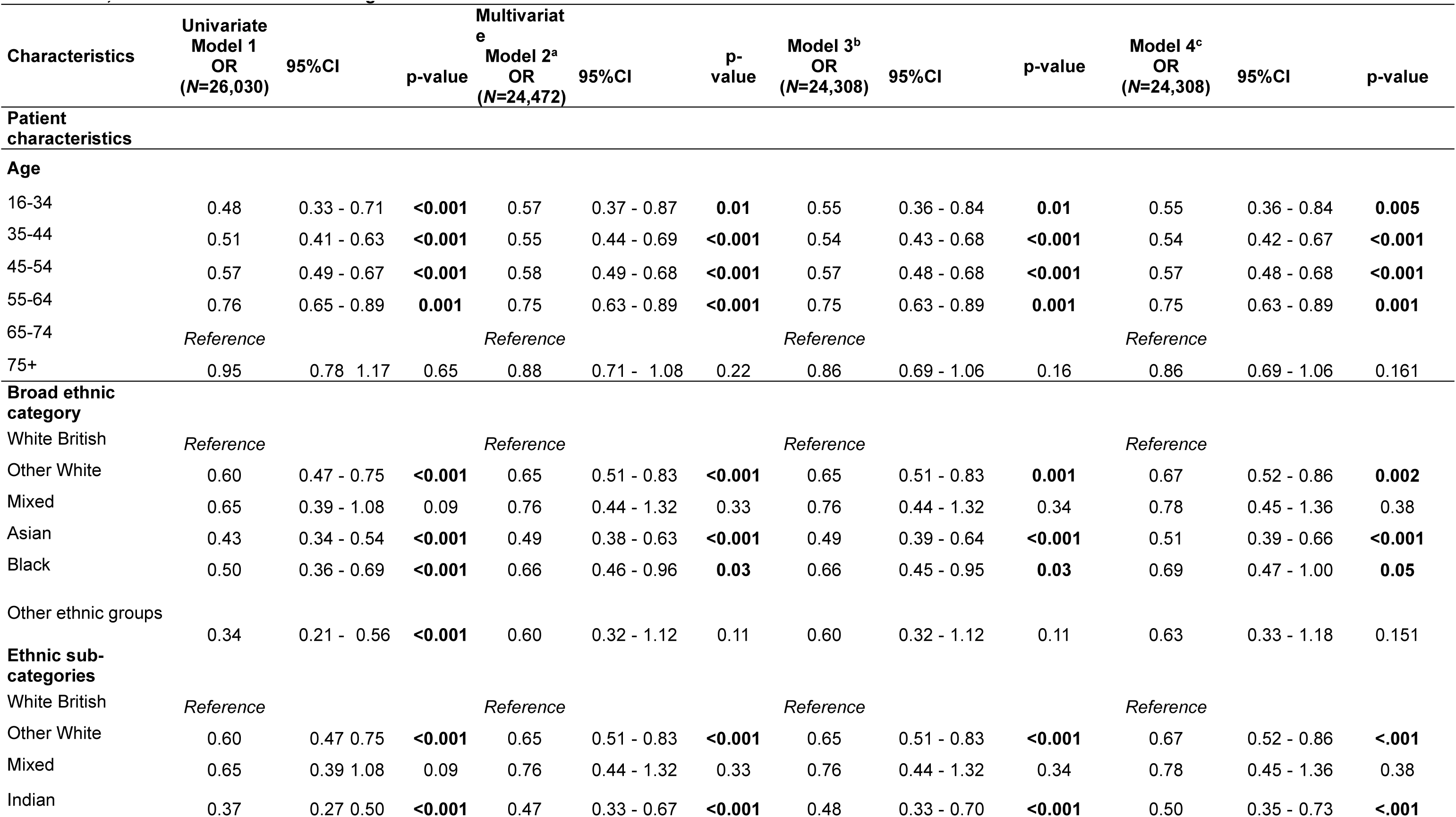
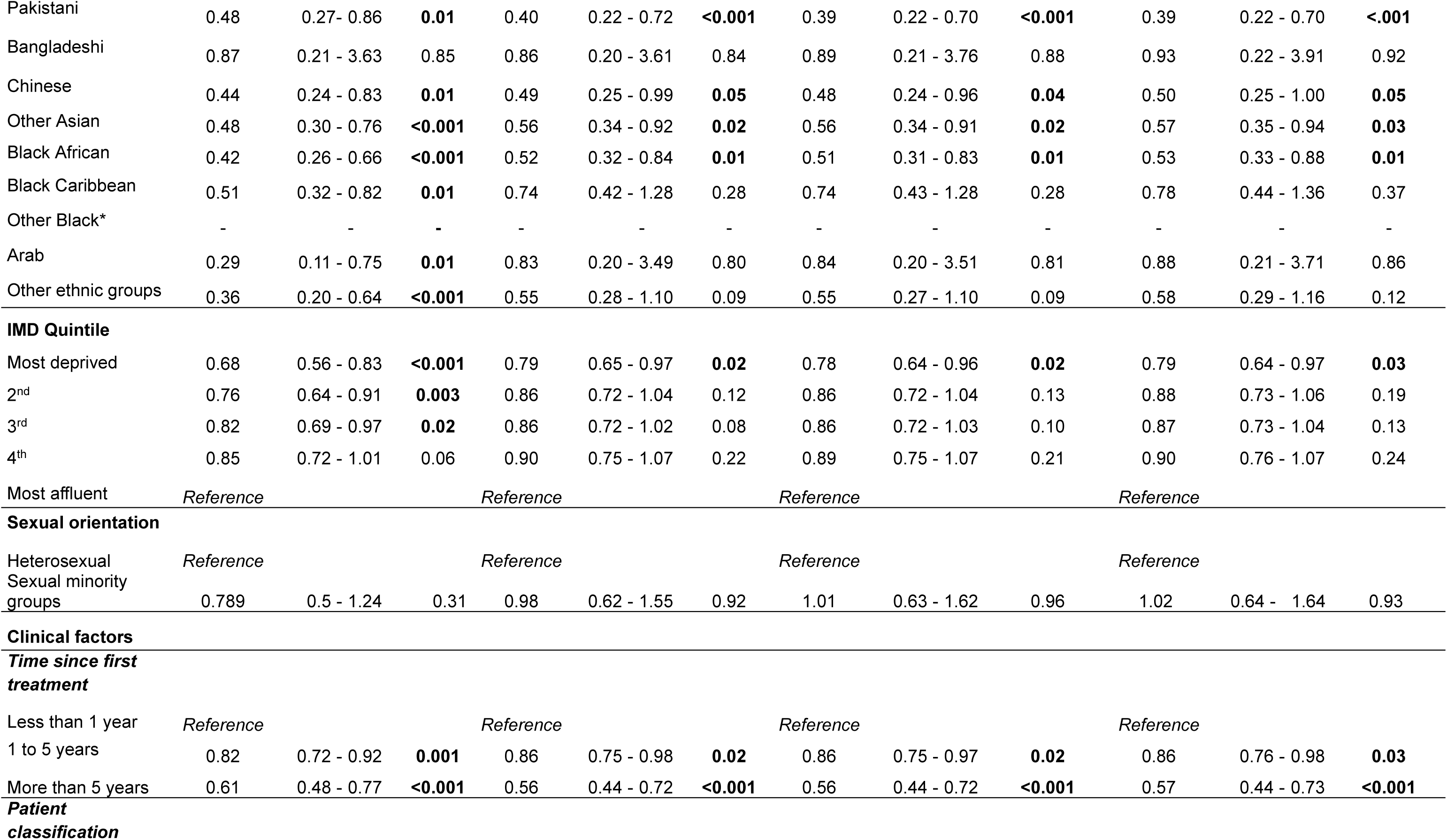
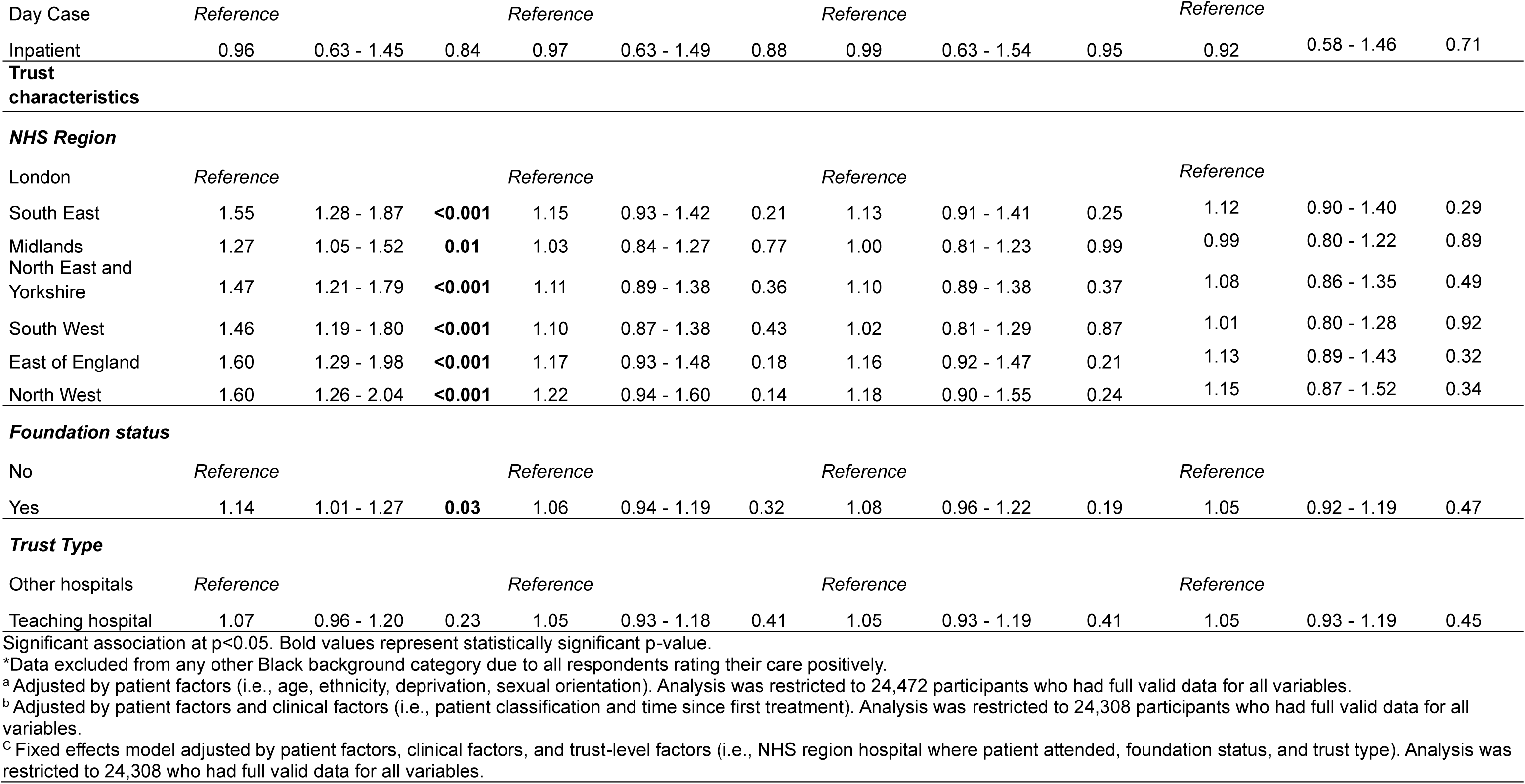
Association between positive rating of overall care experience and demographic characteristics adjusting for patient factors, clinical factors, and trust-level factors using 2017/2018 NCPES.

## Results

In total, 142,889 patients completed the 2017/2018 NCPES surveys. Of these, 26,030 female respondents had a primary breast cancer diagnosis, full valid sociodemographic data (age, ethnicity, IMD), and had responded to Q59 (Figure 1).

### Descriptive statistics

By age, the percentage of women belonging from the most deprived to the most affluent status ranged from 16.5% (age 16-34 year most deprived group) to 28.9% (age 75+ most affluent group). Minority ethnic groups were more deprived compared to White British. In particular, ethnic sub-categories showed that Black African females (41.7%) had the highest deprivation status among all minority ethnic groups. Sexual minority groups were more deprived compared to heterosexual patients.

Table 2 shows the mean and 95% confidence interval of care experience rating by deprivation status intersecting with other dimensions of inequity (age, ethnicity, and sexual orientation). Younger patients (16-44 years) were more likely to rate their breast cancer care less positively than older patients across all deprivation categories. Similarly, minority ethnic groups were more likely to rate their care less positively than White British females across all deprivation categories except for the most affluent group (i.e., Pakistani, Bangladeshi, other Black groups, and Arab rated the care received more positively than White British women in the most affluent group). Arab female patients rated their care as the least positive among the most deprived groups, followed by other Black and Indian. Chinese female patients rated their care as the lowest (least positive) among the second and third most deprived groups.

### Univariate analysis

Table 3 shows the association between positive rating of overall breast cancer care experience and demographic characteristics adjusted by patient, clinical, and trust-level factors. In unadjusted analysis (model 1), younger patients reported substantially less favourable care experience (16-34 vs 65-74 OR_unadj._= 0.48, 95%CI 0.33-0.71). There was strong evidence of variation in overall care experience among ethnic groups. Compared to White British females, this difference was statistically significant for women from Asian (OR_unadj_ =0.43, 95%CI 0.34-0.54), particularly Indian (OR_unadj_ =0.37, 95%CI 0.27-0.50), and Chinese (OR_unadj_ =0.44, 95%CI 0.24-0.83); Black (OR_unadj_ =0.50, 95%CI 0.36-0.69), particularly Black African (OR_unadj_ =0.42, 95%CI 0.26-0.66); and Arab women (OR_unadj_ =0.29, 95%CI 0.11-0.75). There was significant variation between patients of varying deprivation backgrounds (most deprived vs most affluent OR_unadj._ = 0.68, 95%CI 0.56-0.83).

### Multivariate analysis

After adjusting for patient factors (model 2), difference in care experience by age persisted; however, patients aged 35-44 rated their breast cancer care the least positive (OR_adj._= 0.55, 95%CI 0.44, 0.69). Among ethnic groups, Asian (OR_adj._ =0.49, 95%CI 0.38-0.63) were more likely to rate the care experience less favourable compared to White British females. In particular, Pakistani (OR_adj._ =0.40, 95%CI 0.22, 0.72), Indian (OR_adj._= 0.47, 95%CI 0.33, 0.67), and Chinese (OR_adj._= 0.49, 95%CI 0.25, 0.99) females were associated with the lowest proportion reporting their care experience as positive among all ethnic sub-categories. Among Black ethnic groups, Black African female patients rated their breast cancer care less positively than their White British counterparts (OR_adj._= 0.52, 95%CI 0.32, 0.84). Variation in care experience by deprivation was attenuated showing weak evidence for all categories compared to the most affluent group; however, variation between the most deprived compared to most affluent persisted (OR_adj._= 0.79, 95%CI 0.65-0.97). Differences in breast cancer care experience remained for age, ethnicity, and deprivation after adjusting for patient and clinical factors (model 3) and after adding trust factors (model 4). However, in the adjusted analysis, variation in care experience among Arab females was not statistically significant (OR_adj._= 0.88, 95%CI 0.21-3.70) suggesting that their care experience rating was confounded by patient, clinical, and trust factors. No departure from odds-ratio multiplicativity was evident in interaction analysis between ethnicity and deprivation (i.e., no multiplicative interaction on the odds ratio scale).

## Discussion

To our knowledge, this is the first study to examine intersectional differences in care experience among women living with breast cancer in England. Our analysis of the 2017/2018 NCPES surveys demonstrated that there is marked variation in care experience between and within females subgroups, despite evidence suggesting that breast cancer patients often rate their care more favourably than other groups of cancer patients (Bone et al., 2014). This variation remained after adjusting for patient, clinical, and trust factors. These findings suggest that differential in cancer care experience is not because of confounding factors, rather there are real differences in the care experience among females with breast cancer living in England.

In this study, we identified that less favourable care experience was reported predominantly by the younger most deprived groups. Similarly, less positive care experience was predominantly reported by the most deprived minoritised ethnic groups, particularly Asian (Indian) and Black females (Black African). A large body of evidence suggests socioeconomic factors (age, ethnicity, socioeconomic status) are key drivers of inequities in cancer care experience (Alessy et al., 2022). However, guided by the intersectionality framework, we have built a more precise map of inequities in breast cancer care experience. In doing so, we have identified specific social locations where inequities are exacerbated for some groups. It is therefore important that future research focuses on ascertaining the causal processes and findings solutions to tackle inequities in breast cancer care experience.

Consistent with the literature (Alessy et al., 2022, Bone et al., 2014), this research found that younger patients rated their overall care experience less favourably than older women. Some authors suggest this may be explained by the generational phenomenon and older patients’ gratitude bias (this is, older patients rate their care by comparison of previous generations who did not have access to free healthcare and innovations) (Bone et al., 2014, Huang et al., 2019). There are, however, other possible explanations. For instance, evidence from Australia suggests that healthcare professionals’ objectification of women as ‘breast cancer patients’ ignoring their mothering role and the issues of mothering in the context of living with breast cancer influenced young patients’ experience of care (Fisher and O’Connor, 2012). Preserving fertility is a challenge for young breast cancer patients and it exposes them to emotional difficulties, distress, and social pressures to fulfil parenting roles (Dahhan et al., 2021, Goldfarb et al., 2016). Therefore, compared to older patients, young breast cancer patients may rate their overall care experience based on the level of psychosocial support and information they receive from healthcare professionals to deal with both their breast cancer diagnosis and fertility preservation plan. With a diverse and multicultural population living in England, further research should be undertaken to better understand what factors may be driving differential in overall care experience among young breast cancer patients.

Similarly, we identified differences in overall care experience between and within ethnic groups. This is, females from Black and Asian communities rated their breast cancer care experience as less favourable than White British females. Particularly, females from Pakistani, Indian, and Chinese ethnic groups were associated with the lowest proportion reporting their care experience as positive. Other groups who also rated their care less favourably compared to White British women, included women from Black African and other ethnic groups. In population health research, it has been suggested that variation in care experience among ethnic groups may be explained by differences in cultural expectations of care (Bone et al., 2014, Lyratzopoulos et al., 2012). One of the limitations with this explanation is that it imposes responsibility over patients rather than reflecting the potential role the healthcare system may play in driving these inequities, such as cancer services not being tailored and culturally appropriate to meet the needs of the diverse populations they serve. To this end, evidence suggests that social processes and issues of power within the healthcare system (racism, stereotyping, cultural insensitivity) place women from minoritised groups in the UK in more disadvantaged social locations making them more vulnerable to poorer outcomes (NHS RHO, 2022). This situation may explain the difference in the way minoritised groups rate their breast cancer care experience. Institutional oppression is not unique to the UK, with evidence suggesting that embedded racism and discrimination in healthcare institutions is a global issue (Shannon et al., 2022b). Therefore, similar differences in breast cancer experience may occur in other countries. Another possible explanation for the way minoritised breast cancer patients rate their overall care experience is the historical trauma and lack of trust in Western medicine (Mouslim et al., 2020), perception that cancer is a ‘White women’s disease’ (Marcu et al., 2022), cancer fear and fatalism (Vrinten et al., 2016), and cultural factors such as cancer being a taboo (Hirko et al., 2022).

One unexpected finding was that no difference in overall care experience was identified among women with breast cancer from Black Caribbean communities, and all women from other Black ethnic backgrounds rated their overall care experience favourably. In contrast, there was statistically significant differential in care experience between other White female breast cancer patients compared to their White British counterparts. This suggests that belonging to a privileged (White ethnic group) or historically marginalised group (minority ethnic group) does not necessarily transfer into privilege or oppressive experience. Rather, systems of privilege and oppression are not fixed and depend on social location, context, and social processes and can affect individuals and communities in a myriad of ways (Collins, 2015).

A principal strength of this study is the conceptual framework. Guided by intersectionality, we contributed to the literature by identifying the heterogeneity of care experience between and within groups of women living with breast cancer; quantifying inequities in overall care experience; and elucidating the underlying axes of power that might shape the experience of care among women with breast cancer in England. Together with the large national sample size which included 26,030 NCPES respondents, another strength of this analysis was the UK setting (England), where access to breast cancer services is universal. In line with other studies (Lyratzopoulos et al., 2012), our findings suggest that differential in overall breast cancer care experience may be present in other countries with universal healthcare coverage.

A few limitations need to be noted in regarding the present study. First, we were interested to understand how socioeconomic position interacts with other aspects of social identity, recognising the unequal location of women in society (Hill, 2016). For this reason, we only included respondents with a female hospital record. However, we recognise that there are other patients that need attention, including transmasculine breast cancer patients whose particular social location relative to gender, sexual orientation, and socioeconomic position may influence the way they experience cancer care. Likewise, patients with a male hospital record who have been diagnosed with breast cancer may have similar or different care experience that those identified in this study, and also needs attention in future research.

The 2017/2018 NCPES surveys did not collect specific data on education, marital status, or employment. Therefore, socioeconomic position (deprivation quintile) was used as a proxy to ascertain the effect of sociodemographic factors. Similarly, we initially planned to use sexual orientation sub-categories. However, the data collected by the NCPES surveys on sexual orientation are fragmented which led us to collapse this variable into a binary category (heterosexual/sexual minority groups). Evidence shows that sexual minorities are more likely to experience stigma and discrimination in healthcare settings (Kamen et al., 2019), and these experiences are exacerbated when they intersect with race/ethnicity (Greene et al., 2020). This means that it is plausible that sexual minority groups, and particularly sexual minority ethnic groups living with breast cancer in England may rate their cancer experience in different way and this warrants further investigation. In the same vein, we could not investigate the effect of disabilities and comorbidities on care experience among females with breast cancer due to the large proportion of missing data. Therefore, the care experience of these groups relative to axes of power remains unknown (‘intersectional invisibility’(Purdie-Vaughns and Eibach, 2008)). Population health research has identified inequities in care experience among cancer patients living with disability (Tosetti and Kuper, 2023) and comorbidity (Fowler et al., 2020). Therefore, high quality routine data collection is necessary to understand the needs of these often overlooked groups in research (Ginsburg et al., 2023).

Moving beyond the assumption that ethnic groups are homogeneous, we used broad and sub-categories to allow us conducting more granular analyses and to elucidate how social identity intertwines with social contexts and axes of power to influence care experience. However, there are certain limitations with the use of self-reported ethnicity even when it is considered to be ‘gold-standard’ (Saunders et al., 2013). This is because ethnicity is a fluid and a multidimensional concept and therefore, we could not ascertain how a person’s self-reported ethnic group may change in their lifetime. Although we increased the statistical power by using data from two consecutives NCPES surveys, White British females were over-represented. This corroborates previous evidence which suggests that minority ethnic groups are less likely to respond to the NCPES survey, and when they do, they are more likely to rate their care less favourably than do their White British counterparts (Trenchard et al., 2016). These factors could introduce potential selection bias to the NCPES survey and these non-response bias may mask or exacerbate the results, for instance if less satisfied patients may be less inclined to respond (Pinder et al., 2016). We however were not able to ascertain the reasons for non-respondents.

Finally, our analysis offers insights of the overall experience of breast cancer care relative to one NCPES question *Q.59 “Overall, how do you rate your care?”*. It is worth noting that the NCPES survey includes 59-questions covering the whole cancer care pathway. Therefore, it is likely that respondents were at different stages of their treatment, and this may have influenced the way they rated their overall care experience. In addition, there may be nuanced differences in care experience unique to patients living at specific intersectional social locations that could only be understood through qualitative work. To this end, this study is part of larger mixed method research. We plan to conduct further quantitative analyses to explore differential in care experience across the whole breast cancer care pathway to better understand where these differences emerge, and qualitative analysis to understand patients’ perceptions and experiences of care in the context of systems of privilege and discrimination.

## Conclusion

Despite breast cancer consistently receiving higher rates of positive care experience compared to all groups of people living with cancer, we reported marked differences in care experience between and within female subgroups, particularly affecting those living at specific intersection of age, ethnicity and socioeconomic position. This demonstrates the importance of avoiding homogenising groups if we are to better understand inequities in cancer care.

Future quantitative, qualitative, and mixed methods research is required to understand the additional needs of minoritised groups and to ascertain the mechanisms underlying breast cancer inequities. We identified a significant under-representation of minoritised groups including minority ethnic groups, sexual minorities, and people living with comorbidities. Efforts should be made to improve data collection to build a more precise picture of breast cancer inequities and to inform quality improvement plans. Cancer policymakers, commissioners, and healthcare professionals should be cognizant of the heterogeneity of lived experience between and within patients resulting from the intertwined effects of social locations and multiple marginalising factors and how these impact breast cancer care experience. This knowledge should be used to build more inclusive, anti-racist and anti-discriminatory care settings, and to develop intersectional policies and implement tailored interventions and improvement initiatives particularly focused on breast cancer patients at higher risk of vulnerability.

## Ethics

Ethical approval is not required since data is from publicly available secondary sources.

## Authors’ contributions

MEFM: conceptualization, analysis, and preparation of original draft. KLW, AM, and ER: aided in developing the aim and study methods and contributed to the drafting and editing of the manuscript. All authors read and approved the final manuscript.

## Data Availability

All data produced in the present work are contained in the manuscript. Survey data available for non-for-profit research from the UK Data Service (UK Data Service)

https://ukdataservice.ac.uk/

## Acknowledgements

This work has been supported by the University of Surrey Doctoral Studentship Award (TV8330).

## Footnotes

1 Question 69: *‘What is your ethnic group?’*

